# Epidemiological Tools that Predict Partial Herd Immunity to SARS Coronavirus 2

**DOI:** 10.1101/2020.03.25.20043679

**Authors:** Yasuhiko Kamikubo, Atsushi Takahashi

**Affiliations:** Biomedical Data Intelligence, Department of Human Health Sciences, Graduate School of Medicine, Kyoto University, Kyoto, 606-8507, Japan; Department of Physical Therapy, School of Health Science and Social Welfare, and Research Institute of Health and Welfare, Kibi International University, Takahashi, Japan

## Abstract

The outbreak of SARS coronavirus 2 (SARS-CoV-2), which occurred in Wuhan, China in December 2019, has caused a worldwide pandemic of coronavirus disease 2019 (COVID-19). However, there is a lack of epidemiological tools to guide effective public policy development. Here we present epidemiological evidence that SARS-CoV-2 S type exited Wuhan or other epicenters in China earlier than L type and conferred partial resistance to the virus on infected populations. Analysis of regional disparities in incidence has revealed that a sharp decline in influenza epidemics is a useful surrogate indicator for the undocumented spread of SARS-CoV-2. The biggest concern in the world is knowing when herd immunity has been achieved and scheduling a time to regain the living activities of each country. This study provides a useful tool to guide the development of local policies to contain the virus.

---

The pandemic of SARS coronavirus 2 (SARS-CoV-2) ^1-4^ fills the world with fear and confusion, threatening the collapse of health care and the global financial crisis. Flattening the epidemic curve to avoid loss of healthcare capacity is a major global strategy, but has the downside of slowing the achievement of full herd immunity and sacrificing economic activity. Once the herd immunity is completed, the government can confidently decide when to lift the restriction. Therefore, the government must predict trends in the outbreak of SARS-CoV-2. There is no waiting in a pandemic and no time to wait for scientific confirmation. At such times epidemiology exists to determine public health policy first and foremost. Unfortunately, no government knows epidemiological methods for predicting their own SARS-CoV-2 status. Here, we developed the world’s first influenza-based epidemiological method as a useful proxy to detect the spread of SARS-CoV-2 and the establishment of partial herd immunity in countries.

## METHODS

### SOURCES OF DATA

Epidemiological data were obtained from the Worldometer (https://www.worldometers.info/coronavirus/), the COVID Tracking Project (https://covidtracking.com/data/), and websites operated by Su Wei (https://covid-2019.live/), the Infectious Disease Surveillance Center, National Institute of Infectious Diseases, Japan (https://nesid4g.mhlw.go.jp/Hasseidoko/Levelmap/flu/2019_2020/trend.html), the Hokkaido government (http://www.pref.hokkaido.lg.jp/hf/kth/kak/hasseijoukyou.htm), and the World Health Organization (https://app.powerbi.com/view?r=eyJrIjoiZTIxMzAwMzYtZWE4NC00YTU2LWE3MTUtMTI0OGY1ZjQyMWViIiwidCI6ImY2MTBjMGI3LWJkMjQtNGIzOS04MTBiLTNkYzI4MGFmYjU5MCIsImMiOjh9).

### STATISITICAL ANALYSIS

The quantification and modeling of the epidemiological curve was performed under contract with Shin Nippon Biomedical Laboratories, Ltd. (Tokyo, Japan). Analyses of Spearman correlation coefficient were performed with the use of the Statcel4 add-in package (OMS Publishing, Tokorozawa, Japan) for Microsoft Excel.

## RESULTS

A population genetic study of 103 SARS-CoV-2 genomes has identified two viral subtypes designate S and L.^5^ The S type is the ancestral version of the virus. Notably, only 1 out of 27 (3.7%) viral strains isolated from Wuhan was S type, while 10 out of 31 (32.3%) from other places in China and 18 out of 45 (40.0%) outside the country were S type. The L type has accumulated more mutations than the S type and the *orf1ab* replicase/transcriptase gene of L type has a codon that is more preferred for translation than S type, suggesting that the L type replicates and/or transmits faster than the S type. However, it remains to be determined whether the L type is more virulent than the S type.

We examined whether viral subtypes isolated from countries outside China are correlated with patterns of SARS-CoV-2 epidemics. As shown in Table 1, lower ratio of isolated S type is associated with delayed spread of SARS-CoV-2. Moreover, the epidemic seems to be subsiding in areas where S type virus is predominant. These results prompted us to hypothesize that (i) S type SARS-CoV-2 exit Wuhan or other epicenter in China earlier than L type virus without recognition by infectious disease surveillance systems of China and other countries; (ii) the infection by S type induced herd immunity that provides at least partial protection against spread of SARS-CoV-2. Clonal competition between L and S subtypes should have taken place in Wuhan and it is likely that the L type took over the S type because of higher replicative potential and/or faster transmission. Asymptomatic ratio of Japanese nationals evacuated from Wuhan, who are more likely to be infected by the L type, is 30.8% (95% confidence interval: 7.7-53.8%).^6^ In contrast, 51.4% (318/619) of RT-PCR confirmed cases among passengers in the cruise ship Diamond Princess, an outbreak traced to a Hong Kong passenger who is most likely to have S type virus, were asymptomatic (https://www.niid.go.jp/niid/en/2019-ncov-e/9417-covid-dp-fe-02.html). Although the subtypes of the virus remain to be clarified, the higher asymptomatic ratio among cruise ship passengers supports the view that infection by S type is more likely to be asymptomatic than that by L type and easier to avoid recognition by surveillance systems.

**Table 1.**
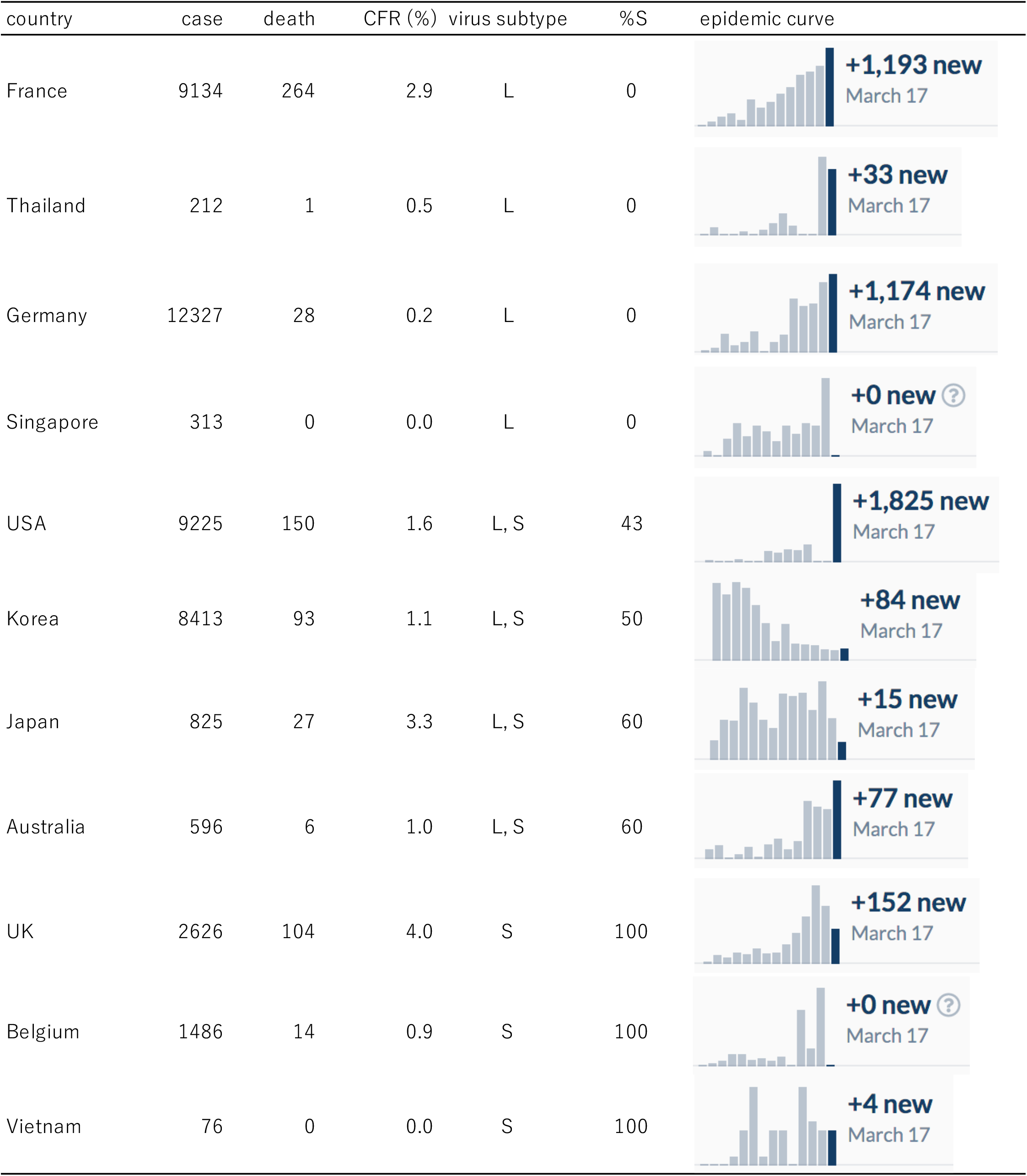
Epidemic Data and Virus Subtypes. Number of cases, the number of death, CFR (%): case fatality rate, virus subtype, %S (the % of S type virus), and epidemic curve are shown according to countries.

| country | case | death | CFR (%) | virus subtype | %S | epidemic curve |
| --- | --- | --- | --- | --- | --- | --- |
| France | 9134 | 264 | 2.9 | L | 0 | **+1,193 new**<br>March 17 |
| Thailand | 212 | 1 | 0.5 | L | 0 | **+33 new**<br>March 17 |
| Germany | 12327 | 28 | 0.2 | L | 0 | **+1,174 new**<br>March 17 |
| Singapore | 313 | 0 | 0.0 | L | 0 | **+0 new ?**<br>March 17 |
| USA | 9225 | 150 | 1.6 | L, S | 43 | **+1,825 new**<br>March 17 |
| Korea | 8413 | 93 | 1.1 | L, S | 50 | **+84 new**<br>March 17 |
| Japan | 825 | 27 | 3.3 | L, S | 60 | **+15 new**<br>March 17 |
| Australia | 596 | 6 | 1.0 | L, S | 60 | **+77 new**<br>March 17 |
| UK | 2626 | 104 | 4.0 | S | 100 | **+152 new**<br>March 17 |
| Belgium | 1486 | 14 | 0.9 | S | 100 | **+0 new ?**<br>March 17 |
| Vietnam | 76 | 0 | 0.0 | S | 100 | **+4 new**<br>March 17 |

Medical doctors in Japan have noticed that influenza epidemic suddenly faded this winter season. Type I interferons induced by S type SARS-CoV-2 may interfere with infection by influenza virus. We therefore performed regional disparity studies comparing influenza warning reports from prefectures in Japan with current positive rate of RT-PCR for SARS-CoV-2. Only 15 prefectures were selected where more than 400 samples were examined because positive rates in other prefectures show Poisson distribution and are not reliable as an indicator of incidence. As shown in Fig. 1, the presence of sharp declines in influenza warning reports correlated with lower incidence of SARS-CoV-2 infection. From early November 2019 to 23 January 2020, about 1,840,000 tourists from China entered Japan, suggesting that unrecognized spread of S type SARS-CoV-2 interfered with influenza epidemic. Interestingly, the timing of decline suggests that S type SARS-CoV-2 spread from 23 December 2019 in Tokyo, Kanagawa, Saitama, and Hiroshima; from 20 January 2020 in Fukuoka and Niigata; from 27 January in Kyoto, Shizuoka, and Ibaraki; and from 2 February in Wakayama. No outbreak has been recognized in Japan before February 2020 and the Chinese government implemented blockades in Wuhan and other cities in Hubei Province on 23 January 2020, suggesting that only S form entered those prefectures.

**Figure 1.**
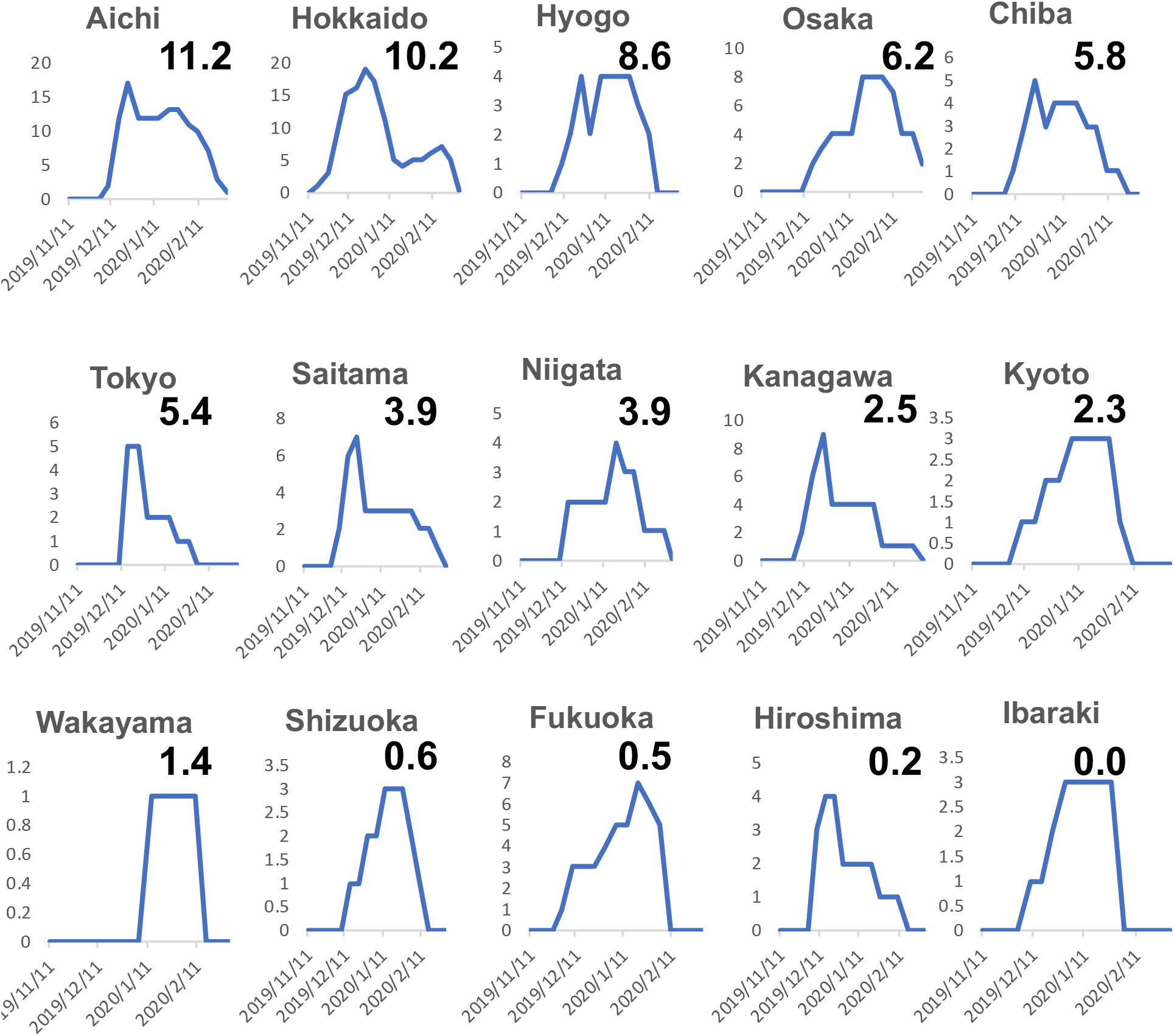
SARS-CoV-2 Trend Curves in Japan Prefectures. The number of influenza warning reports from November 11, 2019 to March 8, 2020 has been plotted. The positive rate (%) of RT-PCR for SARS-CoV-2 as of March 16, 2020 is shown for each prefecture.

The maximum number of infected cases in Japan has been reported in Hokkaido. Fig. 1 suggests that Hokkaido was not or incompletely exposed to S type and the current epidemic is due to the L type. Mathematical modeling has shown that undocumented infections of SARS-CoV-2 were the infection source for most documented cases.^7^ Because children infected with SARS-CoV-2 are asymptomatic^8^ or mildly symptomatic,^4^ we surmise that children are major sources of SARS-CoV-2 spread. Hokkaido is divided into several General Promotion Bureau jurisdictions.

Incidents of influenza-like illness in kindergartens reported from health centers in the jurisdictions were compared with current prevalence of SARS-CoV-2. As shown in Fig. 2, strong negative correlation was observed (Spearman correlation coefficient ρ= -0.73), suggesting that undocumented infection SARS-CoV-2 in Hokkaido suppressed influenza-like illness among small children. Influenza-like illnesses among older children did not reflect the epidemic (ρ= -0.14 in elementary school, 0.07 in junior high school, and -0.04 in high school), possibly because influenza-like symptoms of COVID-19 in these age groups^4^ confounded the analyses.

**Figure 2.**
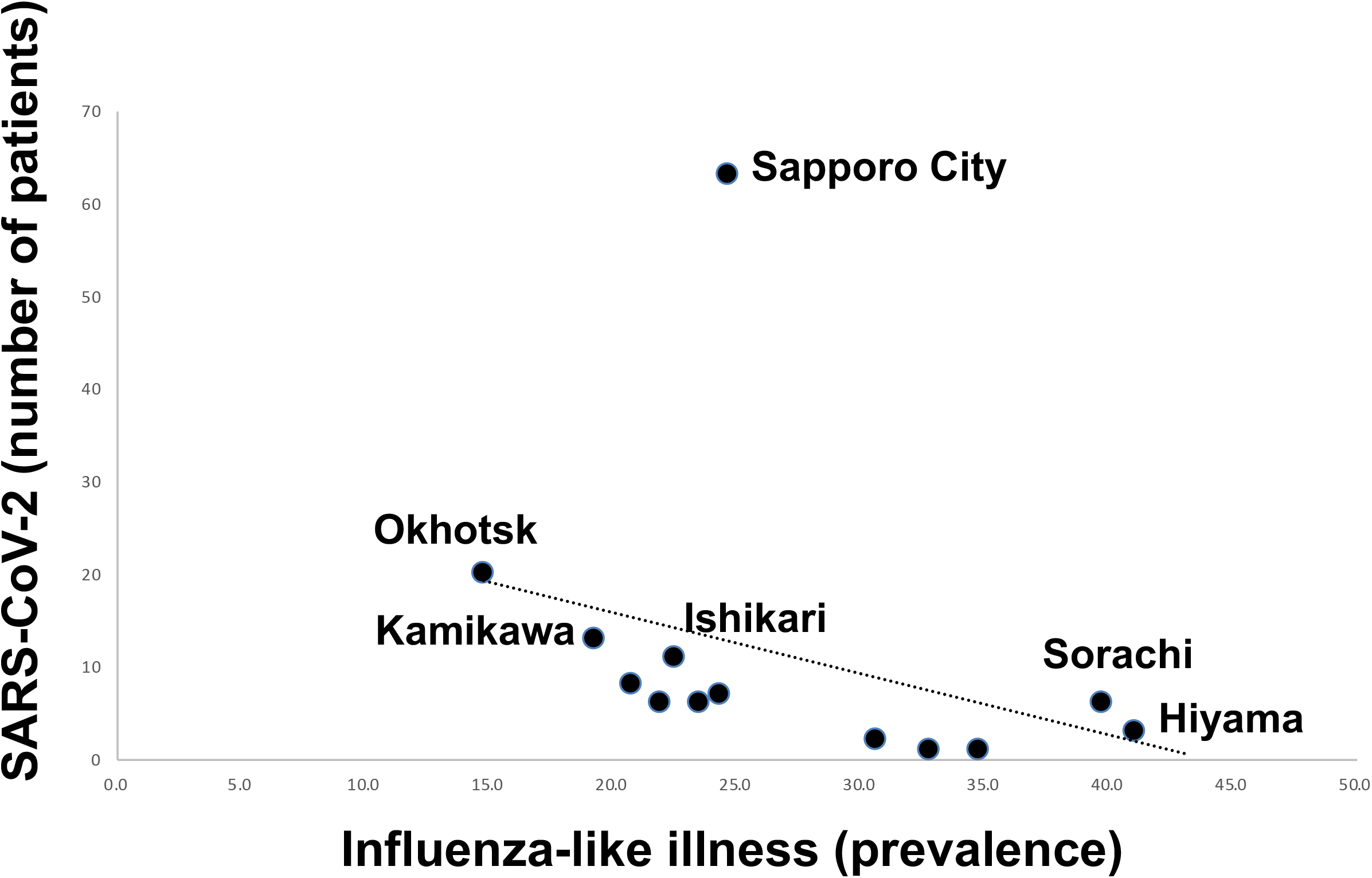
Correlation between Influenza in Kindergartens and SARS-CoV-2 Infection. The correlation diagram shows the prevalence (%) of influenza-like illness in kindergartens in each district in Hokkaido and the number of individuals who were RT-PCR positive for SARS-CoV-2. Spearman correlation coefficient ρ = -0.73.

Europe has become the center of the pandemic. But why mortality rates vary from country to country remains enigmatic. We arranged influenza epidemic curves of European countries in descending order of COVID-19 mortality (Supplementary Figs. 1 and 2). There seems to be a pattern in the curve related to COVID-19 case fatality rates (CFR). Depending on the likelihood of S-type SRS-CoV-2 transmission, a scoring system was developed to model the mortality-related curve patterns. As shown in Fig. 3, this score correlates with the SARS-CoV-2 CFR in each country, implying that the extent of S type SARS-CoV-2 transmission determines the severity of the current infection. This scoring system is useful as a country-specific COVID-19 severity risk score. As shown in Fig. 4, the risk score reveals geographic effects on the spread of the S type SRS-CoV-2.

**Figure 3.**
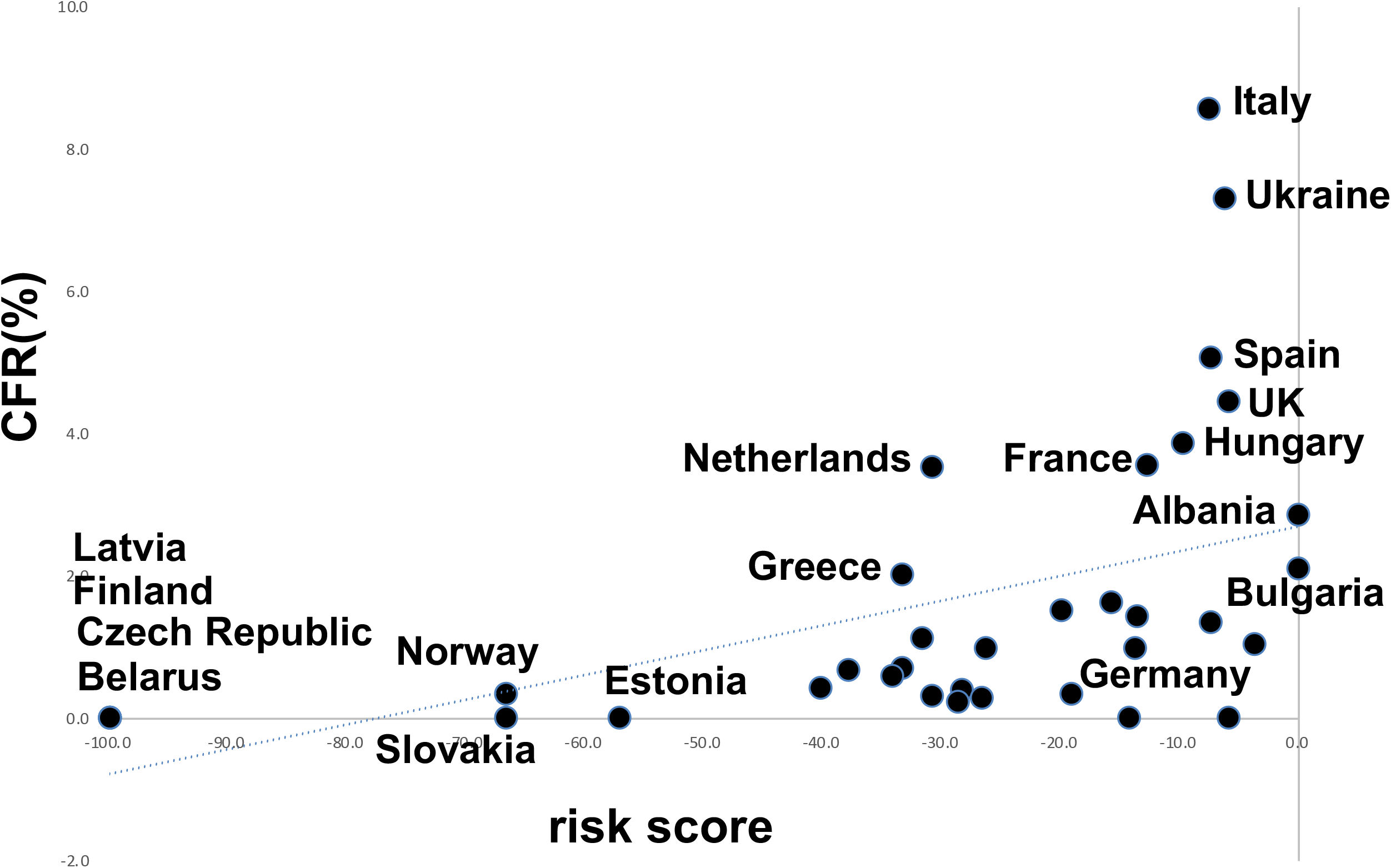
Correlation between the Risk Score and COVID-19 Mortality. Risk scores and case fatality rate (CFR) of COVID-19 in European countries calculated from data in the Worldometer were plotted. Spearman correlation coefficient ρ = 0.65.

**Figure 4.**
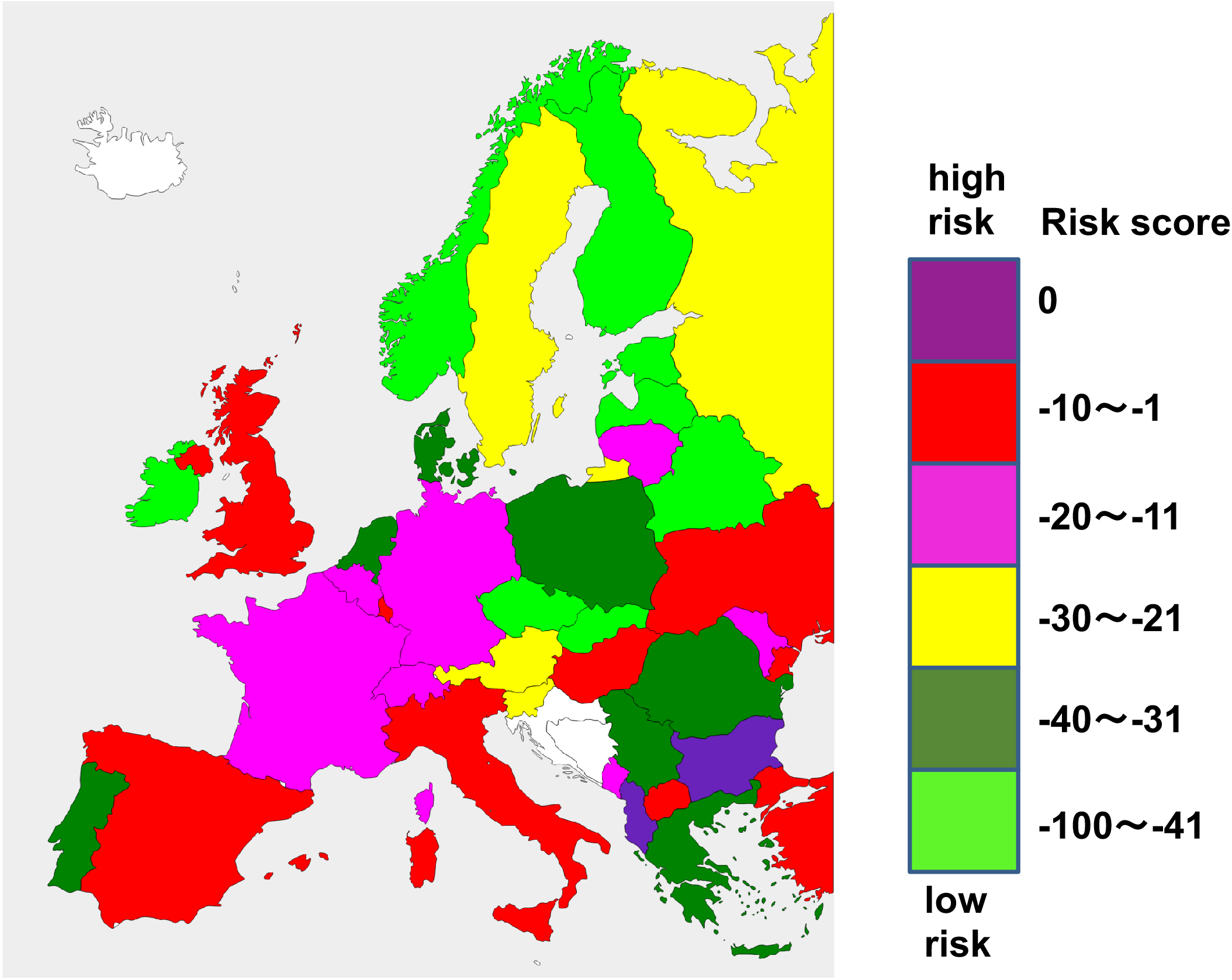
European Map. Distribution of risk scores of various strengths. Data on influenza epidemics were not available for blank areas.

## DISCUSSION

Our epidemiological analyses provided evidence for herd immunity against the S type SARS-CoV-2 that exited Wuhan or other epicenter in China earlier than the L type. Undocumented spread of SARS-CoV-2 can be monitored through influenza surveillance systems based on viral competition. The findings have led to the development of an epidemiological tool that predicts severity of COVID-19 in each region and its application in Europe.

Because the L type is likely to transmit faster than the S type, the basic reproduction number of the L type (*R*_0_^L^) is expected to be higher than the S type (*R*_*0*_^S^). When herd immunity to the S type is established,, the proportion of the population with immunity (*p*_S_) is 1 - 1/*R*_*0*_^S^.^9^ If *R*_*0*_^L^ = 2.2 and *R*_*0*_ ^S^ = 2.17, which are plausible values,^10^ *p*_L_ = 1 - 1/*R*_*0*_ ^L^ = 0.55 (55%) of the population will be infected when a non-immune population is exposed to the L type. When a population with herd immunity to the S type is exposed to the L type, *p*_L_ - *p*_S_ = 1/*R*_*0*_^S^ – 1/ *R*_0_^L^ = 0.006 (0.6%) will be infected, consistent with the positive rates of SARS-CoV-2 RT-PCR in some Japan prefectures (Fig, 1). If 80% of a population is exposed to the S type and then the whole population is exposed to the L type, *p*_L_ – 0.8 × *p*_S_ = 1 - 1/*R*_*0*_^L^ – 0.8 (1 - 1/ *R*_0_^L^) = 0.114 (11.4%) is infected, consistent with the positive rates observed in Aichi and Hokkaido in Japan (Fig. 1). Thus, the spread of S type before the L type confers partial herd immunity to the L type.

Until March 9, 2020, the Japanese government did not restrict entry of tourists from areas of China outside Hubei Province which might have unintentionally allowed selective influx of S form to Japan. S type influx into Ireland and the Balkans may be due to travel restrictions in those countries restricted to infected individuals. In contrast to other areas in Japan, epidemic is ongoing in Nagoya City in Aichi prefecture and Sapporo City in Hokkaido, where the spread of the S form may have been incomplete as discussed above. In the United States of America and most European countries, travel from across China was restricted from the beginning of February, blocking the inflow of the S form from China. There is deep concern that serious outbreaks of SARS-CoV-2 in the United States and Europe may have been caused by the spread of the L form preceding the S form, by which up to 55% of the population may be infected as calculate above.

Although traditional public health outbreak response tactics implemented in China have been shown to be effective,^7^ they incur massive costs such as economic losses.^11^ Conversely, herd immunity policy will face substantial mortality among high risk population.^12,13^ We propose that a policy that expects herd immunity are dangerous in the areas indicated in brown, red, and magenta in Fig. 4. In areas marked in yellow, policies that balance social closure and population immunity are desirable. In the areas marked in green, in contrast, no social isolation may be required except for individuals over 60 and at high risk, such as those with diabetes, coronary heart disease, high blood pressure, chronic obstructive pulmonary disease, cerebrovascular disease, and cancer.^12,13^ Thus, with the use of the new epidemiological tool, governments may be able to predict SARS-CoV-2 morbidity and mortality in their countries and use them in public health policies. Applying this scoring system to small geographies should allow for finer-grained policy creation.

Quantitative analysis for SARS-CoV-2-specific IgM and IgG antibodies in the blood of healthy persons may reveal herd immunity against S type SARS-CoV-2. Such seroepidemiological data will predict the effectiveness and risk of herd immunity policies. Detection of “S type SARS-CoV-2 herd immunity” might also help schedule a timing to lift travel restrictions, event restraints, closure of schools, social distancing, and contact precautions. However, collection of sera and analyses require significant logistic effort, time, and cost,^6^ which developing countries may not be able to cover. Moreover, to determine the normal range of the test, the test values in unaffected individuals must be determined. For diseases with many subclinical infections, such as SARS-CoV-2, it may be difficult to determine the cutoff value,^9^ especially if the subclinical infection is prevalent in the majority of the population. Meanwhile, quick epidemiological tools should be useful for policy making that balances herd immunity approaches with containment policies. The approach used in this study will lay the foundation for further developing epidemiological tools to support political decisions on infection control.

## Data Availability

Data are available on request.

This work was supported by Grant-in-Aid for Scientific Research (KAKENHI; 17H03597 and 16K14632) from the Japan Society for the Promotion of Science.

We thank Hiroyuki Asakura for suggesting epidemiological analyses of COVID-19, Eitaro Ogawa for collecting samples, Toshio Hattori and Hiroaki Kitano for suggesting the possibility of viral competition, Katsumi Kitagawa and Makoto Naruke for discussions.

## Author contribution

Y.K. and A.T. conceived and designed the study. Y.K. performed the sample collection.

A.T. performed the epidemiological and statistical analyses. Y.K. and A.T. wrote the paper.

## Competing interests

The authors declare no competing interests.

## Notes

### Competing Interest Statement

The authors have declared no competing interest.

